# Not only practicing but suffering bullying is correlated with alcohol, tobacco and drugs use results of the Brazilian National School Health Survey (PeNSE 2019)

**DOI:** 10.1101/2023.12.06.23299408

**Authors:** Giuliana Perrotte, Marjorie Mastellaro Baruzzi, João Maurício Castaldelli-Maia

## Abstract

**Background:** Bullying and the use of psychoactive substances are prevalent conditions among adolescents that appear to have some connection. However, there is no consensus in the literature regarding the association between being a victim of bullying and using psychoactive substances. Moreover, most analyses of this correlation have taken place in developed countries, possibly not reflecting the reality in Latin America.

**Objective:** This cross-sectional study investigated the association between the use of psychoactive substances and involvement in bullying situations in a representative sample of Brazilian adolescents.

**Methods:** We used data from the 2019 National School Health Survey (Pesquisa Nacional de Saúde do Escolar), analyzing 123,261 questionnaires from youths aged 13 to 17. Questions about experiencing or engaging in bullying, lifetime use, and recent use (in the last 30 days) of tobacco, hookah, e-cigarettes, other tobacco products, alcohol, and illicit drugs were considered. Responses on the recent use (in the last 30 days) of the following substances were also analyzed: clove cigarettes, straw cigarettes, marijuana, and crack. Multivariate logistic regression models were used to determine correlations, and odds ratios (OR) were calculated.

**Results:** Participation in bullying situations, whether as a victim or perpetrator, increases the likelihood of using tobacco cigarette, hookah, e-cigarettes, clove cigarettes, straw cigarettes, alcohol, illicit drugs, marijuana, and crack. The non-involved-in-bullying group has a lower chance of using any analyzed substance, followed by the group that is only a victim. Those with the highest likelihood of use are individuals who engage in bullying exclusively, followed by those who experience both situations.

**Conclusion:** In Brazil, being a victim of bullying is associated with the use of various psychoactive substances. Our results align with findings from Latin America, partially differing from studies in developed countries, highlighting the influence of location in understanding these risk associations.

## 1. INTRODUCTION

Bullying and the use of psychoactive substances are significant mental health issues among children and adolescents worldwide. The prevalence of youths affected by perpetration or victimization of bullying globally ranges from 10% to 65% (1), and the worldwide rate of adolescents using psychoactive substances is approximately 25% for alcohol, 20% for tobacco, and 5% for marijuana (2). There are studies investigating the association between these two behaviors in their various facets.

Bullying is characterized by recurrent intentional and aggressive behavior against a victim in a situation where there is a real or perceived imbalance of power. Furthermore, victims feel vulnerable and powerless to defend themselves, occurring most commonly in a school environment (3). Bullying behaviors can be physical (hitting, kicking, pinching), verbal (name-calling, swearing, teasing), moral (defaming, slandering, discriminating), sexual (abusing, harassing, insinuating), psychological (intimidating, threatening, stalking), material (stealing, robbing, damaging belongings), and virtual (teasing, discriminating, defaming through the internet and mobile devices) (4). Studies have associated the experience of victimization with low self-esteem, physical and emotional symptoms, anxiety, fear, headaches, enuresis, school avoidance, depression, suicidal ideation, and suicide, among others (5). Moreover, the consequences of being a perpetrator or victim of bullying may persist throughout the school years and into adulthood (6).

Similarly to bullying, the use of psychoactive substances can compromise the physical, emotional, and social development of adolescents. Research links this behavior to involvement in accidents and conflicts, difficulties in academic and professional performance, engaging in risky sexual behavior (without the use of condoms) with multiple partners, low self-esteem, and is associated with conditions such as depression, anxiety, minor psychiatric disorders, conduct disorders, and early mortality (7,8). Furthermore, it is crucial to emphasize that drug consumption is more detrimental among children and adolescents, as this is the period when significant transformations occur in the central nervous system, given the maturation and preparation for the new functions and activities of adulthood (9,10). In Brazil, the rate of substance use among adolescents is even higher compared to the global average, making the issue even more relevant. 60.5% of adolescents had already used alcohol, and 25.5% had used some illicit drug (11).

Moreover, there are possible causes for the association between bullying and the use of psychoactive substances. Adolescents involved in bullying are often accompanied by deviant peers, where substance use is more likely to occur. This may lead adolescents who did not use substances before to initiate them as a form of adaptation, thereby reinforcing their ties with deviant peers (12). Another possible explanation includes a potentially shared personality basis for bullying and substance use, providing greater impulsivity that generates both aggressive behavior and the use of psychoactive substances (13).

A systematic literature review (7) including 40 studies aimed to identify how involvement in bullying situations and the use of psychoactive substances in adolescence are associated. The article also establishes an association between bullying perpetration and substance use; however, it fails to define any relationship between victimization by bullying and substance use. It presents 12 cross-sectional studies that found a correlation between victimization by bullying and the use of psychoactive substances and 7 cross-sectional studies that did not find this association, highlighting the uncertainty in the existing literature on this issue.

The majority of studies conducted on this topic have originated from wealthy countries in Europe and North America, not reflecting the scenario in Latin America, even though research indicates that bullying may be even more prevalent in low- and middle-income countries (14,15). Additionally, the few studies conducted in Latin America (16–18) do not analyze a wide range of substances, restricting their focus to tobacco, alcohol, and cannabis only. This study aims not only to generate more data to address the uncertainty regarding the association between bullying and substance use in adolescence but also to investigate this association with less discussed psychoactive substances, such as crack and E-cigarettes.

## 2. METHODS

### 2.1. Study Design and Sample

The data for this study were extracted from the National School Health Survey conducted in 2019 by the Brazilian Institute of Geography and Statistics (IBGE) in Brazil (19). The 2019 National School Health Survey (Pesquisa Nacional de Saúde do Escolar de 2019, PeNSE 2019) was a cross-sectional study covering various themes, including general information, nutrition, physical activity, cigarette use, alcohol consumption, use of other drugs, home and school situations, mental health, hygiene and oral health, safety, use of health services, and body image.

The PeNSE 2019 sample was designed to estimate population parameters (proportions or prevalences) for students aged 13 to 17 years from both public and private schools at different geographic levels: Brazil, Major Regions, Federal Units, Capital Cities, and the Federal District. Data were collected from approximately 4,361 schools across 1,288 Brazilian municipalities. Schools were selected in each allocation stratum with probabilities proportional to their size, measured by the number of classes reported in the 2017 School Census.

Sample size calculation was performed to dimension the sample. Participants who did not agree to participate in the survey, did not provide age information, were younger than 13 or older than 17, were excluded from the sample in this study. Subsequently, adolescents who did not respond to questionnaires on psychoactive substance use or bullying were also excluded. Therefore, initially, 35,500 individuals were removed based on refusal to participate and age criteria, and 1,153 were considered missing values due to non-response to psychoactive substance and bullying questionnaires. The total sample size for this article was 123,261 adolescents.

### 2.2. Ethical Aspects

PeNSE 2019 was approved by the National Commission for Ethics in Research (CONEP), which regulates health research involving human subjects. The aim was to safeguard ethical principles and ensure the confidentiality of information about the interviewed adolescents, as stated in CONEP Opinion No. 3,249,268, dated April 8, 2019 (19).

### 2.3. Measures

The use of psychoactive substances was considered the dependent variable in this study. The analysis covered lifetime use of the following substances: tobacco, hookah, e-cigarette, other tobacco products, alcohol, and illicit drugs. Recent use (in the last 30 days) of the following substances was also examined: tobacco, hookah, e-cigarette, Bali cigarette, straw cigarette, other tobacco products, alcohol, illicit drugs, marijuana, and crack. In the original questionnaire, recent substance use is quantified in terms of the number of times in the last month. However, in our study, we considered “Yes” for substance use one time or more in the last month and “No” for no substance use in the last month.

Bullying was considered an independent variable. Participants were categorized based on their responses to the questions: “In the last 30 days, how many times have any of your schoolmates teased, mocked, ridiculed, intimidated, or made fun of you to the point that you felt hurt, bothered, upset, offended, or humiliated?” and “In the last 30 days, have you teased, ridiculed, mocked, intimidated, or made fun of any of your schoolmates to the point that they felt hurt, bothered, upset, offended, or humiliated?” To form the categories, adolescents who were bullied at least once in the last month were considered bullying victims, and those who answered “Yes” to the second question were considered bullying perpetrators. Our categories were “Not a victim or perpetrator of bullying (No-No),” “Victim only, not a perpetrator of bullying (Victim Only),” “Perpetrator of bullying only, not a victim (Perpetrator Only),” and “Victim and perpetrator of bullying (Victim and Perpetrator).”

Other independent variables considered in this article were gender (boys, girls, no response), age (category 13-15 years and category 16-17 years), and ethnicity (white, no response, and non-white, including black, brown, yellow, indigenous).

### 2.4. Analyses

Descriptive analyses of the dataset were performed, considering the number of individuals and the percentage of the sample engaging in each studied behavior. Furthermore, multivariate logistic regression models were conducted to examine the relationship between bullying and each dependent variable. This analytical method was chosen as it is the most suitable statistical technique for handling dichotomous dependent variables and multiple non-binary independent variables (20).

Subsequently, an adjusted model was conducted, considering gender, ethnicity, and age using the survey module, which determines post-stratification weights considering primarily geographic strata, the administrative dependence of schools (public or private), and their size in terms of the number of classes (19). All analyses were carried out using R version 4.3.0. Results are presented with Odds Ratios and a 95% Confidence Interval. Statistical significance was considered when the p-value was less than 0.05.

## 3. RESULTS

The majority of the population is female (50.83%), non-white (59.86%), and aged 13 to 15 years (65.89%). The “Perpetrator Only” group has a higher percentage of male population compared to the other groups (67.55%). Descriptive analysis results of the sample characteristics are presented in Table 1.

**TABLE 1:** Population characteristics divided by groups.

|  |  | No-No |  | Victim Only |  | Perpetrator Only |  | Victim and Perpetrator |  | Total |  | P value |
| --- | --- | --- | --- | --- | --- | --- | --- | --- | --- | --- | --- | --- |
| Gender | Male | 34976 | 50,64% | 15970 | 41,17% | 3815 | 67,55% | 5562 | 57,03% | 60323 | 48,94% | p < 0,01 |
|  | Female | 33940 | 49,14% | 22731 | 58,61% | 1820 | 32,22% | 4162 | 42,67% | 62653 | 50,83% |  |
|  | Missing | 158 | 0,23% | 85 | 0,22% | 13 | 0,23% | 29 | 0,30% | 285 | 0,23% |  |
| Etnia | White | 26973 | 39,05% | 14565 | 37,55% | 1984 | 35,13% | 3497 | 35,86% | 47019 | 38,15% | p < 0,01 |
|  | Non-white | 40742 | 58,98% | 23464 | 60,50% | 3539 | 62,66% | 6036 | 61,89% | 73781 | 59,86% |  |
|  | Missing | 1359 | 1,97% | 757 | 1,95% | 125 | 2,21% | 220 | 2,26% | 2461 | 2,00% |  |
| Age | 13 - 15 | 44065 | 63,79% | 26736 | 68,93% | 3636 | 64,38% | 6784 | 69,56% | 81221 | 65,89% | p < 0,01 |
|  | 16 - 17 | 25009 | 36,21% | 12050 | 31,07% | 2012 | 35,62% | 2969 | 30,44% | 42040 | 34,11% |  |
| Total |  | 69074 |  | 38786 |  | 5648 |  | 9753 |  | 123261 |  |  |

Groups: "No-No" (Not a victim or perpetrator of bullying); " Victim Only" (Is a victim but not a perpetrator of bullying); " Perpetrator Only" (Is a perpetrator of bullying but not a victim); " Victim and Perpetrator" (Is both a victim and perpetrator of bullying).

The most experimented drugs by this sample are alcohol (62.15%), hookah (21.75%), and cigarettes (20.04%), respectively. It can be observed that the percentage of substance use progressively increases for all substances included in the study in the groups “No-No,” “Victim Only,” “Victim and Perpetrator,” and “Perpetrator Only,” respectively. Descriptive analysis of alcohol and drug use according to the categories “No-No,” “Victim Only,” “Perpetrator Only,” and “Victim and Perpetrator” is reported in Table 2.

**TABLE 2:**
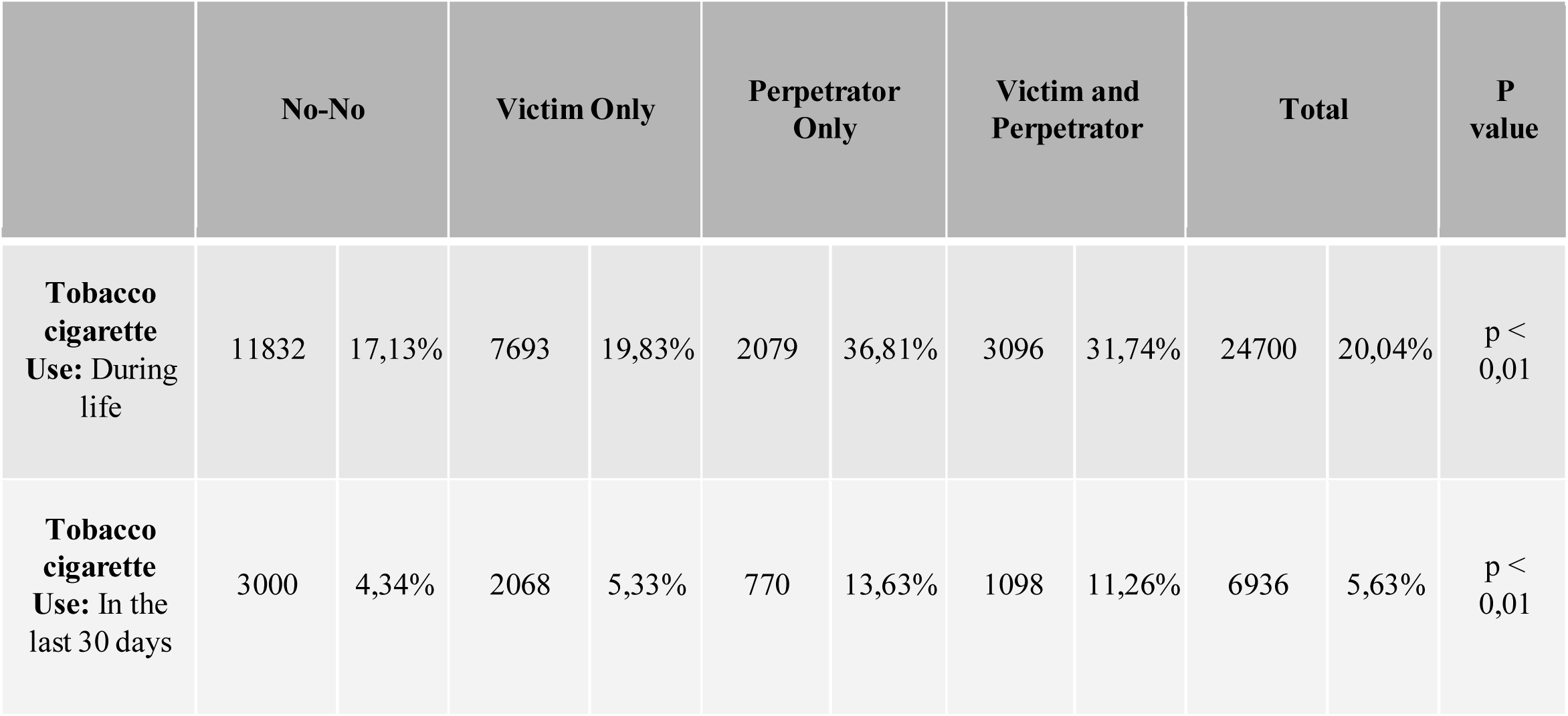
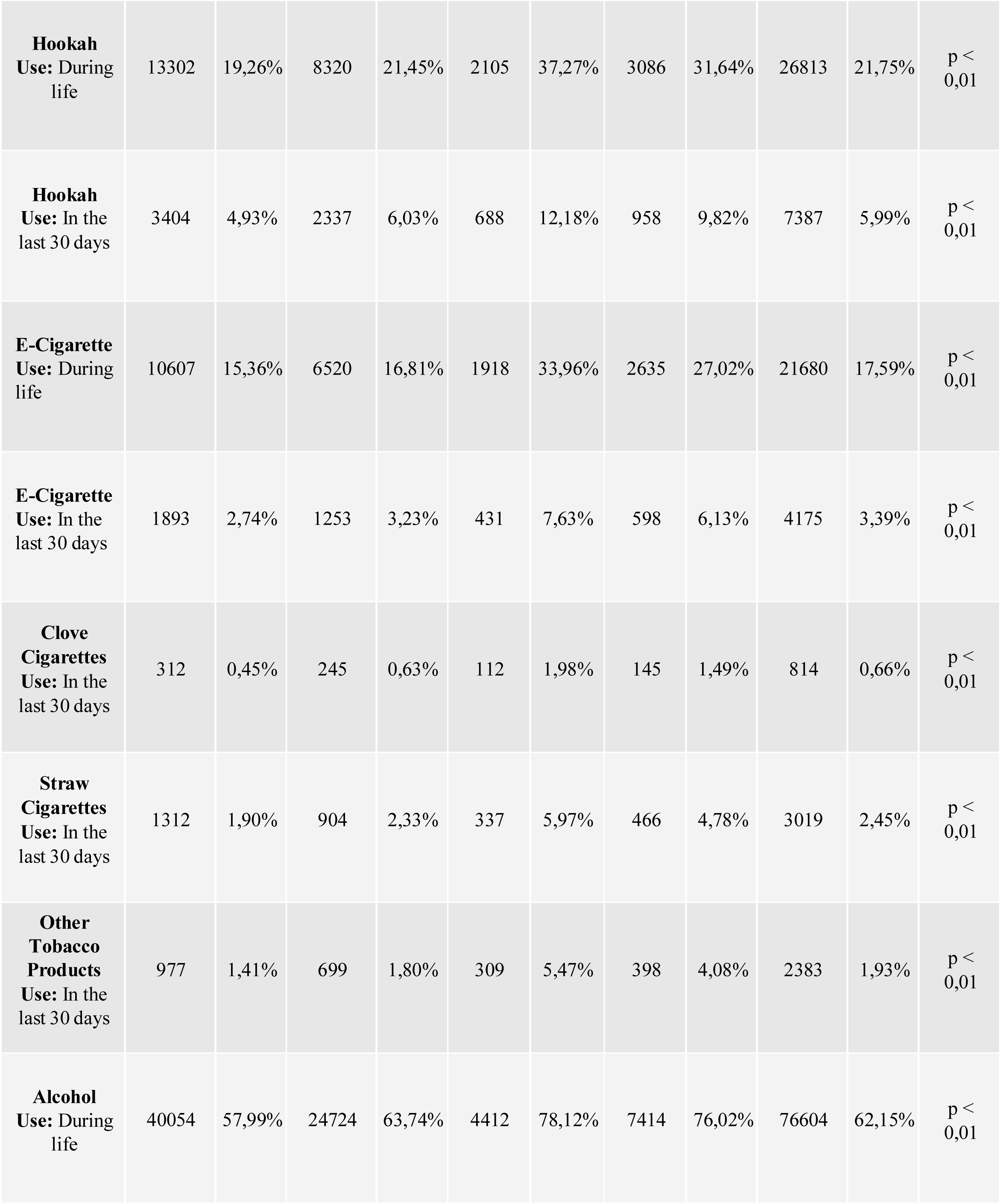
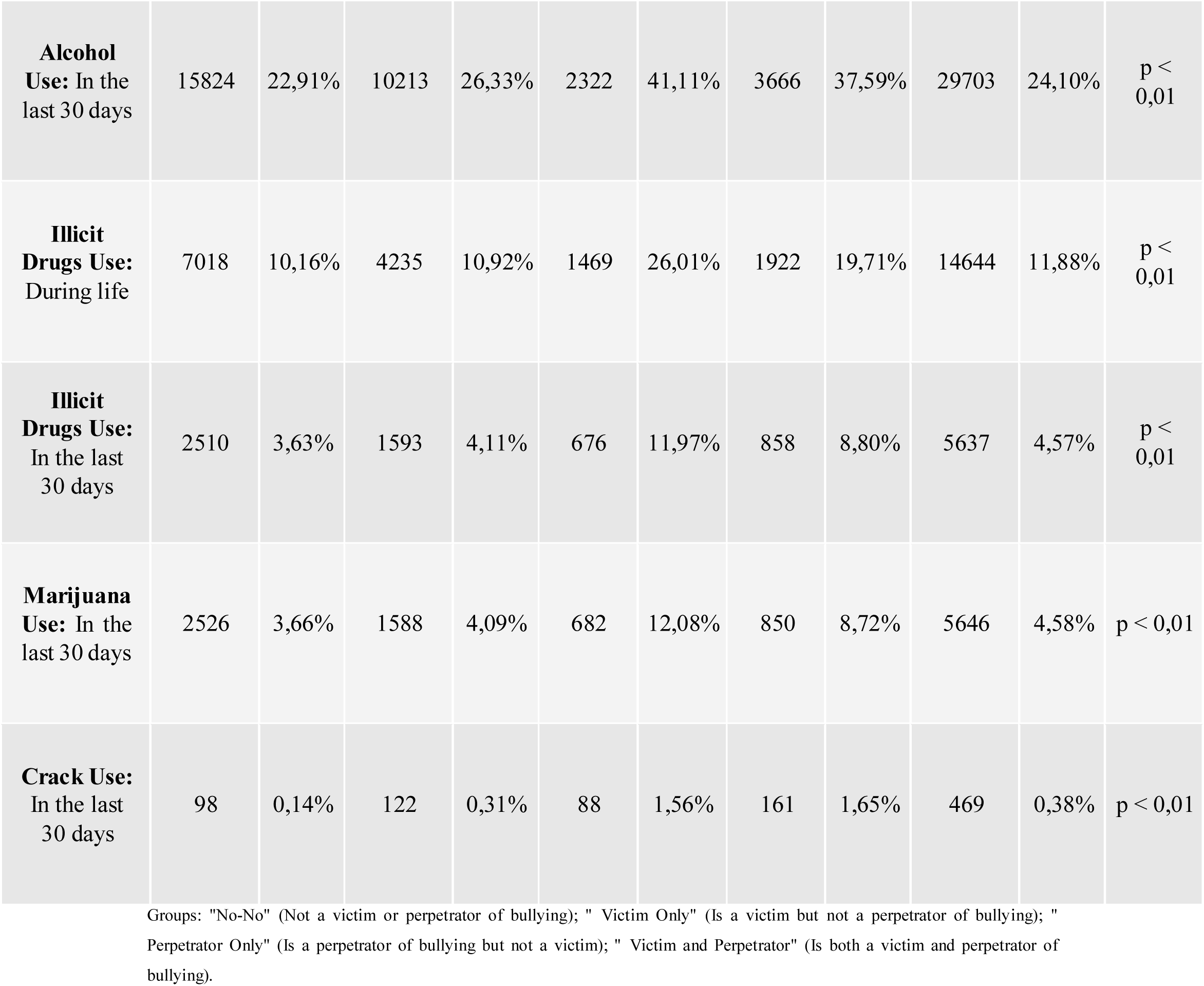
Description of psychoactive substance use in the sample divided by groups.

The data demonstrate that being in the group that is a victim and/or perpetrator of bullying significantly increases the relative risk of using all substances included in the study. The “No-No” group is the subset of the sample with the lowest chance of using any substance in the study. Next, the group most protected against substance use is the “Victim Only” group. The groups with the highest chance are “Perpetrator Only” and “Victim and Perpetrator,” with the highest chance belonging to the “Perpetrator Only” group. Table 3 clarifies the results of adjusted logistic regression models for gender, age, and ethnicity.

**TABLE 3:**
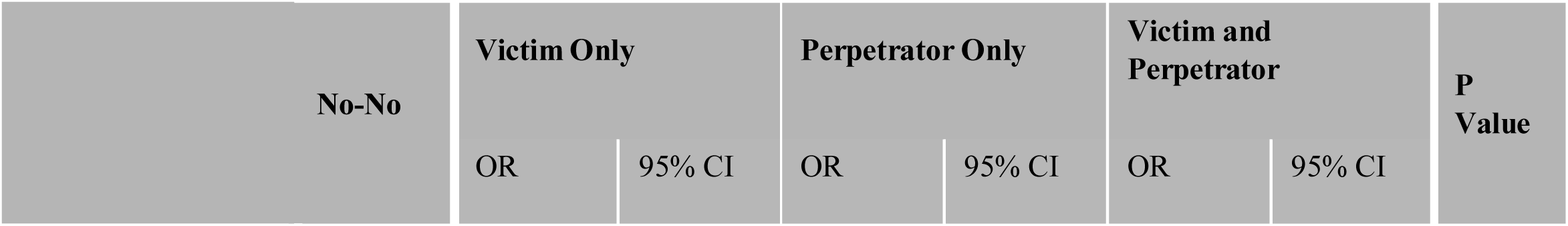
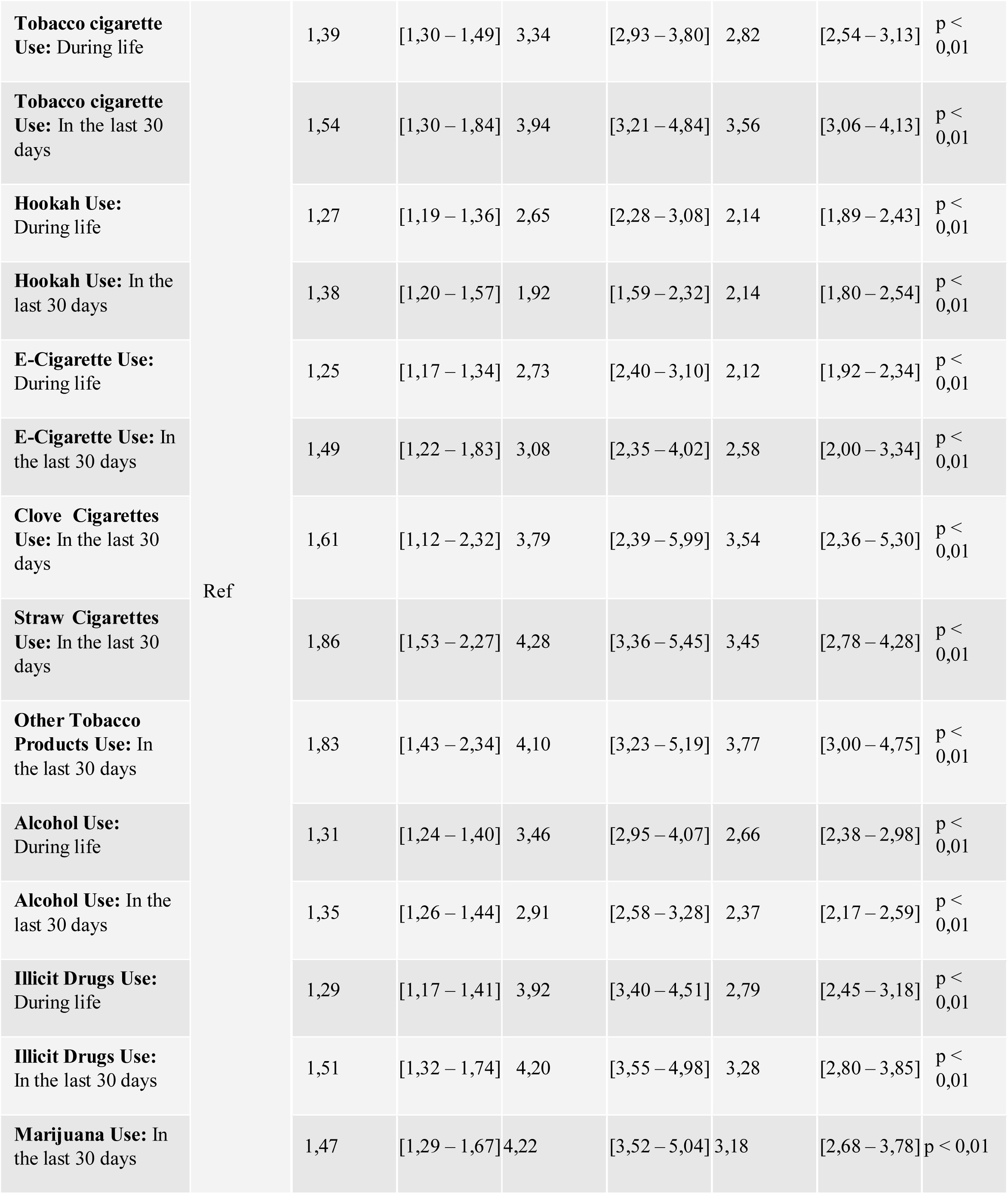
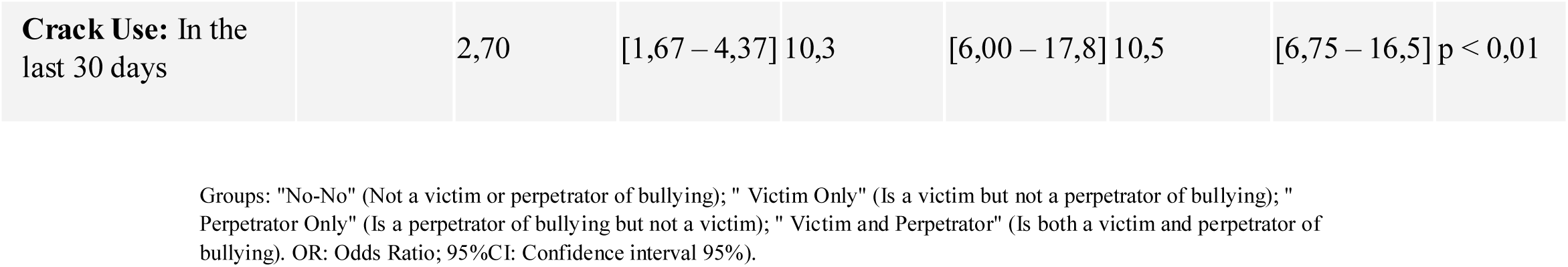
Logistic regression analysis between bullying and the use of psychoactive substances.

### 3.1 Tobacco

Our logistic regression model showed that being a victim and/or perpetrator of bullying has a statistically significant correlation with smoking tobacco cigarettes, both in the “ever in life” and “in the last 30 days” categories. The “Victim Only” group has a lower correlation compared to the “Perpetrator Only” group (“ever in life”: OR: 1.39; 95% CI [1.30 – 1.49]; “in the last 30 days”: OR: 1.54; 95% CI [1.30 – 1.84]). The group with the highest correlation is the “Perpetrator Only” group (“ever in life”: OR: 3.34; 95% CI [2.93 – 3.80]; “in the last 30 days”: OR: 3.94; 95% CI [3.21 – 4.84]). The “Victim and Perpetrator” group also has a higher chance of smoking tobacco cigarettes compared to the “No-No” group (“ever in life”: OR: 2.82; 95% CI [2.54 – 3.13]; “in the last 30 days”: OR: 3.56; 95% CI [3.06 – 4.13]).

### 3.2 Hookah

In our study, being a victim and/or perpetrator of bullying also had a statistically significant correlation with smoking hookah, both in the “ever in life” and “in the last 30 days” categories. The “Victim Only” group again had the lowest correlation compared to the “Perpetrator Only” group (“ever in life”: OR: 1.27; 95% CI [1.19 – 1.36]; “in the last 30 days”: OR: 1.38; 95% CI [1.20 – 1.57]). The group with the highest correlation again was the “Perpetrator Only” group (“ever in life”: OR: 2.65; 95% CI [2.28 – 3.08]; “in the last 30 days”: OR: 1.92; 95% CI [1.59 – 2.32]). The “Victim and Perpetrator” group also has a higher chance of smoking hookah compared to the “No-No” group (“ever in life”: OR: 2.14; 95% CI [1.89 – 2.43]; “in the last 30 days”: OR: 2.14; 95% CI [1.80 – 2.54]).

### 3.3 E-Cigarette

Another substance that has an increased chance of being consumed, both in the “ever in life” and “in the last 30 days” categories, if the school-going adolescent is a victim and/or perpetrator of bullying is the E-Cigarette. Again, the increase in chances is progressively higher in the groups “Victim Only” (“ever in life”: OR: 1.25; 95% CI [1.17 – 1.34]; “in the last 30 days”: OR: 1.49; 95% CI [1.22 – 1.83]), “Victim and Perpetrator” (“ever in life”: OR: 2.12; 95% CI [1.92 – 2.34]; “in the last 30 days”: OR: 2.58; 95% CI [2.00 – 3.34]), and “Perpetrator Only” (“ever in life”: OR: 2.73; 95% CI [2.40 – 3.10]; “in the last 30 days”: OR: 3.08; 95% CI [2.35 – 4.02]), respectively.

### 3.4 Other Tobacco Products

This analysis also evaluated the correlation of bullying with the use of clove cigarettes, straw cigarettes, and other tobacco products in the last 30 days. Once again, the pattern of increased correlation of substance use in the groups “Victim Only” (clove cigarettes: OR: 1.61; 95% CI [1.12 – 2.32]; straw cigarettes: OR: 1.86; 95% CI [1.53 – 2.27]; other tobacco products: OR: 1.83; 95% CI [1.43 – 2.34]), “Victim and Perpetrator” (clove cigarettes: OR: 3.54; 95% CI [2.36 – 5.30]; straw cigarettes: OR: 3.45; 95% CI [2.78 – 4.28]; other tobacco products: OR: 3.77; 95% CI [3.00 – 4.75]), and “Perpetrator Only” (clove cigarettes: OR: 3.79; 95% CI [2.39 – 5.99]; straw cigarettes: OR: 4.28; 95% CI [3.36 – 5.45]; other tobacco products: OR: 4.10; 95% CI [3.23 – 5.19]) is followed, respectively.

### 3.5 Alcohol

The research described here also showed that being a victim and/or perpetrator of bullying has a statistically significant correlation with alcohol consumption, both in the “ever in life” and “in the last 30 days” categories. The “Victim Only” group has a lower correlation compared to the “Perpetrator Only” group (“ever in life”: OR: 1.31; 95% CI [1.24 – 1.40]; “in the last 30 days”: OR: 1.35; 95% CI [1.26 – 1.44]). The group with the highest correlation is the “Perpetrator Only” group (“ever in life”: OR: 3.46; 95% CI [2.95 – 4.07]; “in the last 30 days”: OR: 2.91; 95% CI [2.58 – 3.28]). The “Victim and Perpetrator” group also has a higher chance of alcohol consumption compared to the “No-No” group (“ever in life”: OR: 2.66; 95% CI [2.38 – 2.98]; “in the last 30 days”: OR: 2.37; 95% CI [2.17 – 2.59]).

### 3.6 Illicit Drugs

Illicit drugs also have an increased chance of being consumed, both “ever in life” and “in the last 30 days,” if the school-going adolescent is a victim and/or perpetrator of bullying. Again, the increase in chances is progressively higher in the groups “Victim Only” (“ever in life”: OR: 1.29; 95% CI [1.17 – 1.41]; “in the last 30 days”: OR: 1.51; 95% CI [1.32 – 1.74”]), “Victim and Perpetrator” (“ever in life”: OR: 2.79; 95% CI [2.45 – 3.18]; “in the last 30 days”: OR: 3.28; 95% CI [2.80 – 3.85”]), and “Perpetrator Only” (“ever in life”: OR: 3.92; 95% CI [3.40 – 4.51]; “in the last 30 days”: OR: 4.20; 95% CI [3.55 – 4.98”]), respectively.

### 3.7 Marijuana

According to the analysis described here, being a perpetrator and/or victim of bullying also increases the chance that the adolescent has used marijuana in the last 30 days. Again, the least affected group is “Perpetrator Only” (OR: 1.47; 95% CI [1.29 – 1.67]), and the most affected group is “Perpetrator Only” (OR: 4.22; 95% CI [3.52 – 5.04]), with the “Victim and Perpetrator” group (OR: 3.18; 95% CI [2.68 – 3.78]) being intermediate.

### 3.8 Crack

Among the psychoactive substances analyzed in this research, crack cocaine shows the highest increase in the chance of use when one is a victim and/or perpetrator of bullying. The magnitude of the odds ratio increase follows the same logic as the other substances studied, with the “Victim Only” group being the most protected (OR: 2.7; 95% CI [1.67 – 4.37]); the “Victim and Perpetrator” group intermediate (OR: 10.5; 95% CI [6.75 – 16.5]); and the “Perpetrator Only” group having the highest odds ratio (OR: 10.3; 95% CI [6.00 – 17.8]) when compared to the “No-No” group.

## 4. DISCUSSION

This cross-sectional study with a large representative sample of Brazilian school adolescents aged 13 to 17 showed that being a victim and/or perpetrator of bullying increases the chance of young people using psychoactive substances such as tobacco, hookah, e-cigarettes, clove cigarettes, straw cigarettes, other tobacco products, alcohol, illicit drugs, marijuana, and crack. This finding is important for drawing attention to this correlation, which is poorly understood in the literature.

A recent literature review (21) including 23 studies with adolescents analyzed the correlation between perpetrating bullying and the use of psychoactive substances. The results were similar to ours, suggesting that adolescent perpetrators of bullying are more likely to be involved in the use of alcohol, tobacco, and illicit drugs. A Canadian cross-sectional study (22) including 64,174 school adolescents showed that victimization by bullying was associated with an increased chance of using all types of drugs included in the study, including alcohol, marijuana, and cocaine, aligning with the results described here. Another Canadian cross-sectional study (23) with 49,543 adolescent students showed that victims of bullying were significantly more likely to use an electronic cigarette, a finding also present in our analysis. A large North American study (24) (N=74,059) also showed that being a victim of bullying increases the chance of young people using cigarettes, alcohol, or marijuana, similar to what we discovered.

Studies conducted in Latin American countries also show findings consistent with this study. A Brazilian study (18) involving 102,301 students aged 13 to 18 found a positive correlation between the use of tobacco, alcohol, and cannabis and being a victim of bullying. A similar connection was found in an Argentine study (25) with 1,328 students aged 13 to 15, associating victimization by bullying with a higher chance of alcohol use. In Chile, a similar connection was also found in a representative sample of adolescent students (N=36,687) (26), showing that being a victim of bullying significantly increases the chance of tobacco and alcohol use.

However, there are divergent data regarding the risk of psychoactive substance use among bullying victims, especially in samples from developed countries. Rivers et al. (27) analyzed 2002 young British students in a cross-sectional study and did not find a correlation between being a victim of bullying and the use of psychoactive substances. Forster et al. (28), in a North American longitudinal study involving 1167 adolescents, found a significant association between being a victim of bullying and tobacco use in the last month; however, they did not find an association between being a victim of bullying and the use of alcohol or marijuana in the last month, differing from our results. Hemphill et al. (29), in a longitudinal study involving 907 Australian adolescents, found no correlation between being bullied and the use of alcohol or marijuana in the last two weeks.

Furthermore, another important aspect is that most studies exclusively analyze victims or perpetrators of bullying, lacking a group that both practices and experiences bullying simultaneously. It is worth noting the articles that include this group. Radliff et al. (30) conducted a cross-sectional analysis of 74,247 American adolescent students, obtaining results very similar to ours. They investigated the use of tobacco, alcohol, and marijuana in the last 30 days and concluded that aggressors have higher prevalences of these outcomes, followed by aggressor-victims, and those not involved have the lowest prevalences, including in relation to victims. Bradshaw et al. (31) also conducted a cross-sectional analysis of 16,302 American adolescent students, investigating the consumption of alcohol, marijuana, and tobacco in the last 30 days. In the results of this study, aggressor-victims appear as the group with the highest likelihood of using all three substances, followed by only aggressors and, lastly, those who were only victims. A study (32) with a large number of participants (N=113,200) associated bullying and alcohol use, finding a correlation between being a perpetrator or victim-perpetrator of bullying and higher alcohol use. However, they did not find a significant association for alcohol use and being only a victim of bullying. It is noteworthy that this study was conducted in European and North American countries, predominantly, which could explain the different findings compared to our analysis.

### 4.1 Implications and Contributions

Our research elucidates the close correlation between bullying and the use of psychoactive substances. This finding holds significant implications for pediatricians, child psychiatrists, psychologists, school professionals, and other professionals involved in adolescent health, who should be trained to investigate and address both situations. Furthermore, considering this data, it can be hypothesized that interventions preventing bullying may reduce the likelihood of substance use among school adolescents, potentially representing a viable public policy to be implemented in schools. However, there is a need for longitudinal studies following the implementation of such measures to confirm the hypothesis.

Amundsen et al. (33) conducted a longitudinal analysis of data from 23 Norwegian schools that had implemented an anti-bullying program, comparing them to two schools without the program. It was found that students in both groups used psychoactive substances at the same frequency; however, students who had experienced the anti-bullying program were less prone to intoxication than students in the control group. Despite the lack of a direct reduction in substance use rates, this analysis suggests that anti-bullying interventions may have positive effects on the sphere of psychoactive substance use among adolescents.

Another recent intervention (34) applied in Brazil assessed the effectiveness of a bullying prevention program in 73 schools. Besides verifying a positive effect against bullying, it also resulted in a indirect decrease in alcohol use among students, supporting the hypothesis that anti-bullying policies may also have a protective effect on psychoactive substance use.

### 4.2 Limitations

The limitations of this study need to be acknowledged. Firstly, as it is a cross-sectional study, establishing causal relationships between being a victim and/or perpetrator of bullying and the use of psychoactive substances is not feasible. It would be valuable for the scientific community to conduct future longitudinal studies with a large number of participants on this topic. However, it is worth noting that the major studies that have investigated this issue so far have also been cross-sectional studies and had a smaller number of participants than the present analysis.

Secondly, it is essential to remember that the questionnaire used in the 2019 PeNSE, the basis of our study, is self-administered and may be subject to incorrect interpretations by participants and memory bias. However, this type of instrument was also used in practically all previous studies. Additionally, the self-administered questionnaire makes adolescents more comfortable in responding to questions that may cause embarrassment, such as those about the use of psychoactive substances.

Thirdly, these data cannot be extrapolated to all Brazilian adolescents. The sample only included adolescents enrolled in schools. However, it is noteworthy that 89.2% of Brazilian adolescents are enrolled in regular schools. Moreover, bullying is a phenomenon that occurs more frequently within the school environment.

## 5. CONCLUSION

The study presented here revealed a correlation between being a victim and/or perpetrator of bullying and the use of psychoactive substances such as tobacco, hookah, e-cigarettes, clove cigarettes, straw cigarettes, other tobacco products, alcohol, illicit drugs, marijuana, and crack. Our results align with findings from Latin America, partially differing from studies in developed countries, emphasizing the influence of location on the understanding of these risk associations. Further research with different methodological designs is needed to delineate causality between the two variables. Considering this data, it can be hypothesized that the implementation of anti-bullying policies in schools brings benefits regarding the use of psychoactive substances among adolescents, making this a valuable discussion.

## Data Availability

All data produced in the present work are contained in the manuscript

## 7. APPENDICES

